# Sex differences in frailty and the impact of frailty on blood pressure control in older adults with hypertension: a multi-center observational study in Vietnam

**DOI:** 10.1101/2024.12.20.24319394

**Authors:** Tan Van Nguyen, Vien Thi Nguyen, Wei Jin Wong, Erkihun Amsalu, Trinh Kim Thi Ngo, Mark Woodward, Tu Ngoc Nguyen

## Abstract

**Background:** Frailty is common in older adults with hypertension and can affect blood pressure (BP) control. Sex differences related to frailty and cardiovascular physiology may contribute to the effective treatment of hypertension.

**Aim:** to examine the prevalence of frailty in older adults with hypertension and its association with uncontrolled BP, with a particular focus on differences by sex.

**Methods:** This study was conducted at the outpatient clinics of two major hospitals in Vietnam from June 2023 to June 2024. Frailty was defined by a Clinical Frailty Scale of ≥4. Uncontrolled BP was defined as systolic BP ≥140 mmHg or diastolic BP ≥90 mmHg, averaged over recordings in the last 6 months. Multivariable logistic regression was applied to identify the association between frailty and uncontrolled BP. The odds ratios (ORs) for uncontrolled BP of each risk factor were estimated by sex, with interaction terms fitted between each risk factor and sex to obtain the women-to-men ratio of ORs (ROR).

**Results:** There were 1038 participants (326 women, 712 men). They had a mean age of 73.3 (SD7.4). The prevalence of frailty was 28.6% in all participants, higher in women (35.3%) than men (25.6%), p=0.001. The overall rate of uncontrolled BP was 26.7%. In women, the frail had a significantly higher rate of uncontrolled BP (33.9%) compared to the non-frail (20.9%), but no significant differences among men (26.4% in the frail vs. 27.5% in the non-frail). The adjusted ORs of frailty on uncontrolled BP were 1.70 (1.00–2.90) in women, 0.84 (0.57–1.25) in men; women-to-men ROR 2.02 (1.04–3.92).

**Conclusion:** In older adults with hypertension, frailty was more common in women and was associated with an increased risk of having uncontrolled BP in women only. These findings highlight the need for sex-specific approaches in managing hypertension in older populations.

## Introduction

High blood pressure is a significant risk factor for the development of cardiovascular disease and a range of other chronic health conditions.^1,2^ The prevalence of hypertension increases with age, being 27% for people aged less than 60, but up to 74% for those aged 80 and older.^3^ Despite efforts to manage hypertension globally, poor blood pressure (BP) control continues to be a challenge, with approximately 20% of people with hypertension achieving optimal BP control.^4,5^ Uncontrolled hypertension can substantially increase the risk of stroke, heart failure, coronary heart disease, peripheral artery disease, renal failure, and dementia.^1,2,6^

In Vietnam, a systematic review in 2019 reported an overall prevalence of hypertension of 21% in adults aged 18 or older.^7^ A recent 2024 community-based study in 2572 adults (mean age 51.5±15.7) found that 36.1% had hypertension and, of individuals receiving antihypertensive treatment, 57.4% had uncontrolled BP (≥140/90 mmHg).^8^ However, there is limited evidence of hypertension in older adults, particularly those with frailty, in Vietnam. The prevalence of frailty is very high in older adults in Vietnam ranging from 11.2% to 21.7% in community-based studies and 18.5% to 54.9% in hospital-based studies.^9–17^ The treatment of hypertension in older adults is particularly challenging due to the complexity of their health needs and the presence of geriatric conditions, such as frailty, multimorbidity, polypharmacy, and cognitive impairment.^18,19^ In frail patients, various factors can contribute to uncontrolled BP, such as low adherence to prescribed medications, changes in pharmacokinetics and pharmacodynamics and clinicians’ concerns about adverse side effects of antihypertensive treatment, such as dizziness and falls.^2,20–22^ In addition, sex differences related to frailty and cardiovascular physiology also contribute to the effective treatment of hypertension. There has been evidence that frailty occurs more frequently, and with greater severity, in women compared to men, which may be attributed to various biological, social, and environmental factors.^23,24^

Therefore, this study aimed to examine the prevalence of frailty in older adults with hypertension and the association between frailty and uncontrolled BP, with a particular focus on sex disparities.

## Methods

This prospective observational study was conducted at the outpatient clinics of two major hospitals in Vietnam (Thong Nhat Hospital in Ho Chi Minh City and University Medical Centre of Ho Chi Minh City) from June 2023 to June 2024. Consecutive patients aged ≥ 60 diagnosed with hypertension who visited the clinics during the study period were recruited. Patients with secondary hypertension, or not being able to provide consent, were excluded from the study.

The study was approved by the Ethics Committees of the University of Medicine and Pharmacy at Ho Chi Minh City (Reference Number 627/HDDD-DHYD, date 26/06/2023). Informed consent was obtained from all participants. The study was conducted in accordance with the Declaration of Helsinki.

Data were collected from patient interviews and medical records. Information obtained included demographic characteristics, lifestyles, height, weight, medical history, duration of having hypertension, frailty, medications, medication adherence, and comorbidities. Body mass index (BMI) was calculated from measured weight and height. Smoking was defined as previous or current smoking. Regular exercise was defined as exercising at least five days per week. Multimorbidity was defined as having any other long term health conditions in addition to hypertension. Polypharmacy was defined as using 5 or more medications on a daily basis.^25^ Medication adherence was assessed using the 10 item Medication Adherence Rating Scale (MARS) developed by Thompson et al.^26^ MARS scores range from 0 to 10, with a higher score denoting better medication adherence. For this study, a MARS score > 5 was defined as adherence to medications.^27^

Mean systolic BP and diastolic BP were calculated from the BP measurements obtained in patients’ medical records in the past 6 months. Uncontrolled BP was defined as a mean systolic BP ≥ 140 mmHg or a mean diastolic BP ≥ 90 mmHg.

Frailty was assessed using the Clinical Frailty Scale (CFS).^28,29^ The CFS score ranges from 1-9, and a score of 4 or greater indicates a frailty status.^28,30^ (Figure 1)

**Figure 1.**
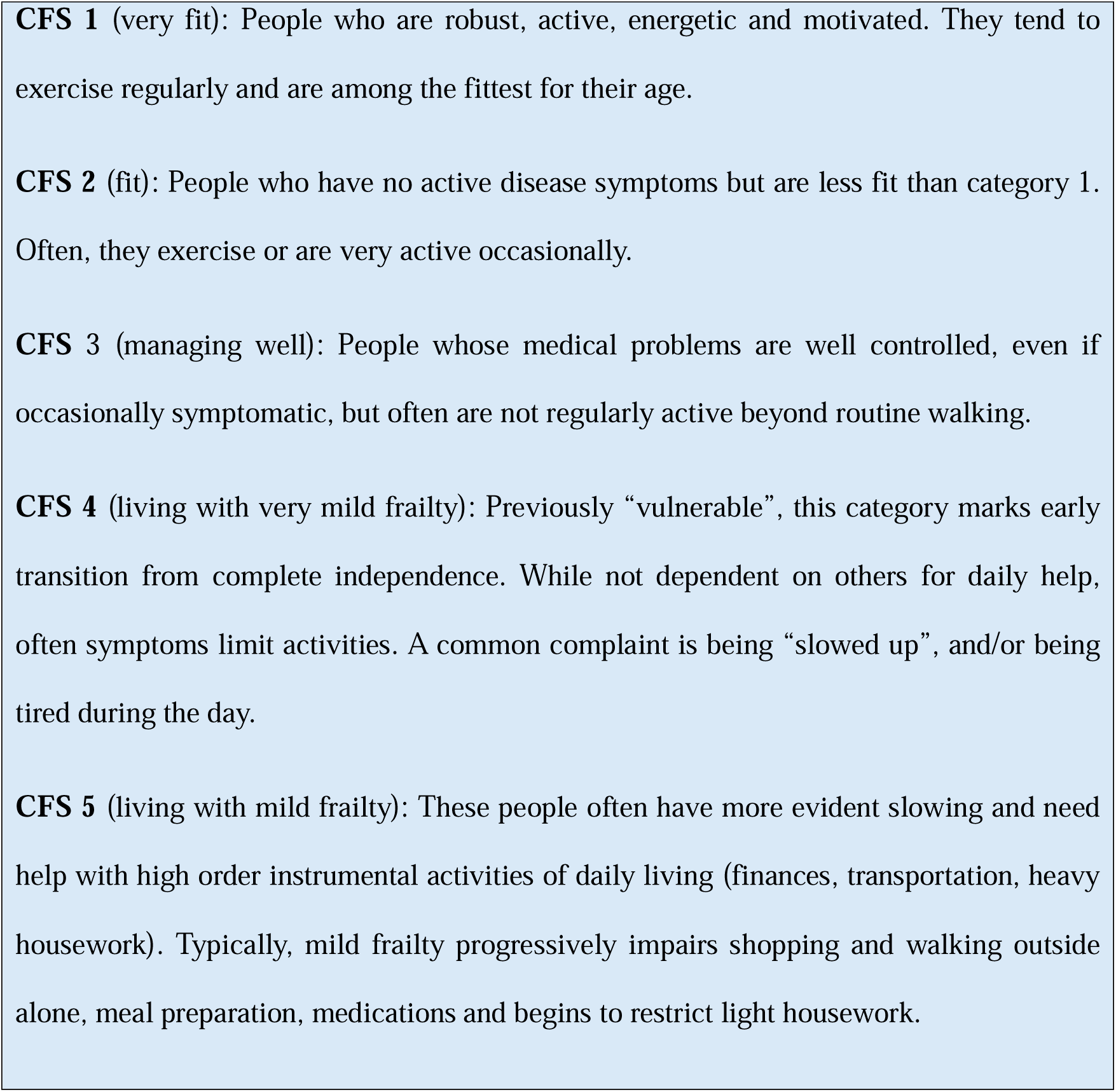
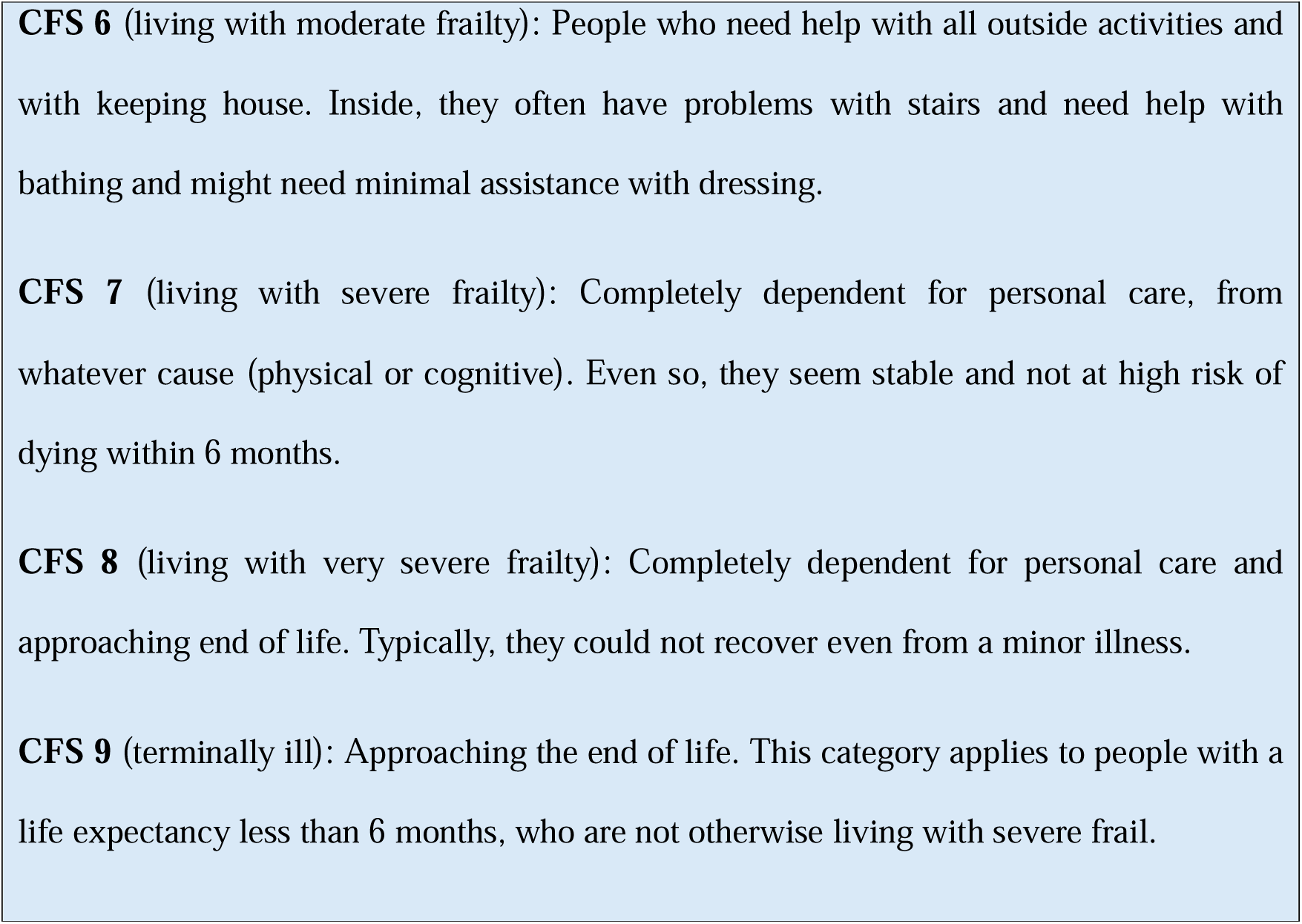
**The Clinical Frailty Scale (CFS)**

Based on the data from Vietnam and from a systematic review of studies worldwide on frailty and hypertension,^9–17,31^ we estimated a prevalence of frailty of 30% in older women and 20% in older men with hypertension. Therefore, at least 784 older adults with hypertension would be needed in this study to detect a significant difference in the prevalence of frailty between women and men (with a power of 90%, 2-sided test, alpha=0.05).

### Statistical analyses

Continuous variables are presented as means and standard deviations (SD), and categorical variables as frequencies and percentages. Comparisons between men and women were assessed using the Chi-square test or Fisher’s exact test for categorical variables and Student’s t-test or Mann-Whitney test for continuous variables.

To identify the association between frailty and uncontrolled BP, multivariable logistic regression models were applied, with frailty as the predictive variable of interest and uncontrolled BP as the outcome variable, adjusting for the covariates. Potential covariates with p-values < 0.05 from unadjusted analyses in either men or women were included in the multivariable logistic regression models. The odds ratios (ORs) for uncontrolled BP of each risk factor were estimated by sex, with interaction terms fitted between each risk factor and sex to obtain the women-to-men ratio of ORs (ROR), based on Woodward’s methodology.^32^ All statistical tests were two-sided and a p value <0.05 was considered as statistical significance.

## Results

There were 1038 participants (326 women and 712 men). They had a mean age of 73.3 (SD 7.4) years (**Table 1**). Women were significantly older than men, with a mean age of 74.3 (SD 7.1) compared to 72.8 (SD 7.5) in men. Coronary heart disease was the most common comorbidity (49.3%), followed by type 2 diabetes (43.8%), and chronic kidney disease (23.0%). Women had significantly higher rates of osteoporosis and falls in the past 12 months. Men had a significantly higher prevalence of coronary heart disease. The percentage receiving combination antihypertensive therapy was significantly higher in men (91.2%) compared to women (86.5%), p=0.022.

**Table 1.** Participant characteristics, stratified by sex.

| Variables | All participants<br>(N=1038) | Women<br>(N=326) | Men<br>(N=712) | P |
| --- | --- | --- | --- | --- |
| Frailty | 297 (28.6) | 115 (35.3) | 182 (25.6) | 0.001 |
| Age (years) | 73.3 (7.4) | 74.3 (7.1) | 72.8 (7.5) | 0.003 |
| Marital status |  |  |  |  |
| Never married | 8 (0.8) | 4 (1.2) | 4 (0.6) | 0.003 |
| Married | 994 (95.8) | 301 (92.3) | 693 (97.3) |  |
| Divorced/separated | 2 (0.2) | 1 (0.3) | 1 (0.1) |  |
| Widow/widower | 34 (3.3) | 20 (6.1) | 14 (2.0) |  |
| Education |  |  |  |  |
| Primary school | 33 (3.2) | 17 (5.2) | 16 (2.2) | <0.001 |
| Secondary school | 193 (18.6) | 86 (26.4) | 107 (15.0) |  |
| High school | 363 (35.0) | 122 (37.4) | 241 (33.8) |  |
| Higher education | 449 (43.3) | 101 (31.0) | 348 (48.9) |  |
| Smoking | 97 (9.3) | 2 (0.6) | 95 (13.3) | <0.001 |
| Regular exercise | 508 (48.9) | 91 (27.9) | 417 (58.6) | <0.001 |
| Body mass index (kg/m<sup>2</sup>) |  |  |  |  |
| <18.50 | 39 (3.8) | 17 (5.2) | 22 (3.1) | 0.196 |
| 18.50-24.99 | 703 (67.7) | 222 (68.1) | 481 (67.6) |  |
| ≥ 25.0 | 296 (28.5) | 87 (26.7) | 209 (29.4) |  |
| History of hospitalization in the past 12 months | 374 (36.0) | 131 (40.2) | 243 (34.1) | 0.059 |
| History of falls in the past 12 months | 89 (8.6) | 45 (13.8) | 44 (6.2) | <0.001 |
| Polypharmacy | 955 (92.2) | 294 (90.5) | 661 (93.0) | 0.163 |
| Medication adherence | 859 (82.8) | 279 (85.6) | 580 (81.5) | 0.103 |
| Multimorbidity | 938 (90.5) | 291 (89.3) | 647 (91.0) | 0.377 |
| Coronary heart disease | 512 (49.3) | 145 (44.5) | 367 (51.5) | 0.035 |
| Type 2 diabetes | 455 (43.8) | 145 (44.5) | 310 (43.5) | 0.777 |
| Chronic kidney disease | 239 (23.0) | 79 (24.2) | 160 (22.5) | 0.532 |
| Osteoarthritis | 214 (20.6) | 64 (19.6) | 150 (21.1) | 0.596 |
| Heart failure | 95 (9.2) | 34 (10.4) | 61 (8.6) | 0.334 |
| Stroke | 85 (8.2) | 28 (8.6) | 57 (8.0) | 0.750 |
| Atrial fibrillation | 67 (6.5) | 22 (6.7) | 45 (6.3) | 0.794 |
| Osteoporosis | 39 (3.8) | 22 (6.7) | 17 (2.4) | <0.001 |
| Chronic obstructive pulmonary disease | 23 (2.2) | 3 (0.9) | 20 (2.8) | 0.055 |
| Antihypertensive therapy |  |  |  |  |
| Monotherapy | 107 (10.3) | 44 (13.5) | 63 (8.8) | 0.022 |
| Combination therapy | 931 (89.7) | 282 (86.5) | 649 (91.2) |  |

Summary statistics shown are n (%), except for age where mean (standard deviation) is given.

In both women and men, the CFS ranged from 2 to 6, with CFS 3 being the most common category (**Figure 2**). The percentages with CFS 5 and CFS 6 were higher in women than in men. The prevalence of frailty overall was 28.6% (297/1038), higher in women (35.3%) than men (25.6%), p=0.001.

**Figure 2.**
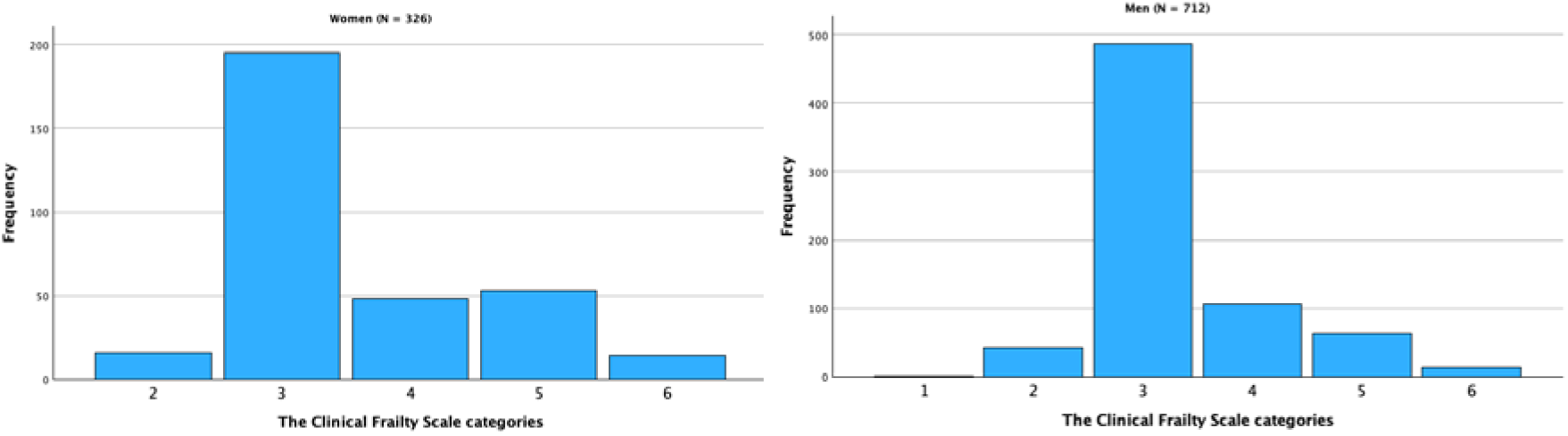
**The distribution of the Clinical Frailty Scale categories in women and men**

Overall, 26.7% of the participants had uncontrolled BP, 29.3% in the frail vs. 25.6% in the non-frail (p=0.229), and 25.5% in women vs. 27.2% in men (p=0.546).

Figure 3 presents the rates of uncontrolled BP stratified by sex and frailty status. Among female participants, the frail had a significantly higher rate of uncontrolled BP (33.9%) compared to the non-frail (20.9%), p=0.010. Among male participants, there was no significant differences in uncontrolled BP rates between the frail (26.4%) and the non-frail (27.5%), p=0.759.

**Figure 3.**
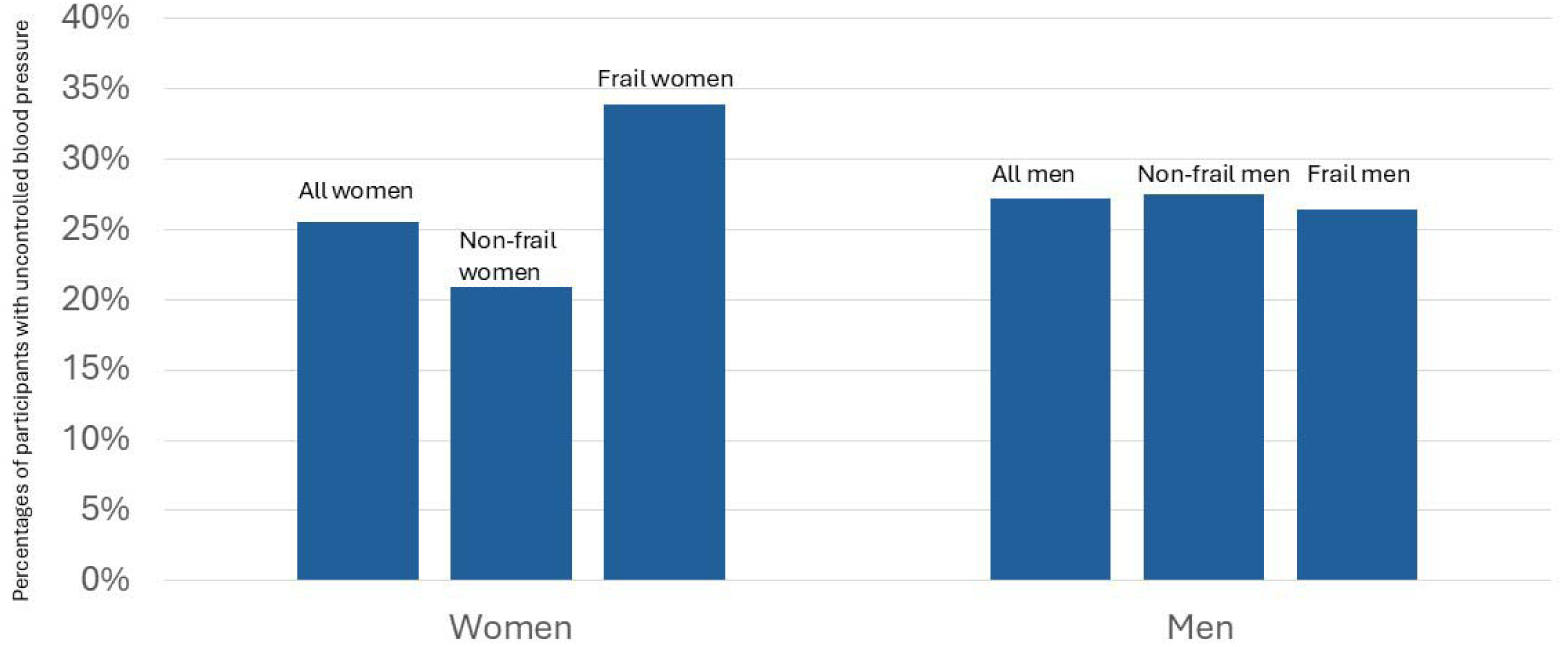
**Uncontrolled hypertension rates by sex and frailty status**

The unadjusted regression analyses of covariates that can be potentially associated with uncontrolled BP are presented in **Table 2**.

**Table 2.** Unadjusted odds ratios (95% confidence intervals) relating potential risk factors to uncontrolled blood pressure in women and in men.

|  | Women (N=326) |  | Men (N=712) |  |
| --- | --- | --- | --- | --- |
|  | Unadjusted OR<br>(95%CI) | P | Unadjusted OR<br>(95%CI) | P |
| Age (years) | 1.03 (0.99 – 1.07) | 0.108 | 0.99 (0.97 – 1.02) | 0.498 |
| Duration of having hypertension (years) | **1.03 (1.00 – 1.06)** | **0.043** | 1.01 (0.99 – 1.03) | 0.402 |
| Marital status (married vs. single/divorced/widowed) | 0.71 (0.29 – 1.70) | 0.437 | 1.42 (0.46 – 4.32) | 0.541 |
| Low education (secondary school or less vs. higher education) | 0.91 (0.53 – 1.57) | 0.738 | 0.88 (0.56 – 1.38) | 0.576 |
| Smoking | 2.95 (0.18 – 47.72) | 0.446 | 0.83 (0.51 – 1.38) | 0.476 |
| Regular exercise | 1.35 (0.79 – 2.32) | 0.278 | 1.36 (0.97 – 1.91) | 0.077 |
| History of hospitalization in the past 12 months | 1.28 (0.77 – 2.11) | 0.345 | 1.03 (0.72 – 1.45) | 0.889 |
| History of falls in the past 12 months | 0.70 (0.32 – 1.52) | 0.367 | 1.75 (0.93 – 3.28) | 0.083 |
| Polypharmacy | 0.99 (0.42 – 2.30) | 0.971 | 0.78 (0.42 – 1.44) | 0.424 |
| Multimorbidity | 0.99 (0.44 – 2.20) | 0.971 | 1.14 (0.63 – 2.06) | 0.667 |
| Coronary heart disease | 0.77 (0.47 – 1.28) | 0.317 | 0.89 (0.64 – 1.24) | 0.501 |
| Type 2 diabetes | 1.22 (0.74 – 2.02) | 0.431 | 1.02 (0.73 – 1.42) | 0.928 |
| Chronic kidney disease | 1.28 (0.73 – 2.25) | 0.392 | 1.10 (0.75 – 1.63) | 0.628 |
| Osteoarthritis | 1.07 (0.58 – 2.00) | 0.821 | 0.67 (0.44 – 1.03) | 0.068 |
| Heart failure | 0.89 (0.39 – 2.05) | 0.785 | 0.86 (0.47 – 1.58) | 0.626 |
| Stroke | 0.97 (0.40 – 2.38) | 0.953 | 1.05 (0.57 – 1.91) | 0.880 |
| Atrial fibrillation | 0.85 (0.30 – 2.39) | 0.761 | 0.86 (0.43 – 1.73) | 0.663 |
| Osteoporosis | 1.11 (0.42 – 2.93) | 0.840 | 1.12 (0.39 – 3.21) | 0.839 |
| Chronic obstructive pulmonary disease | N/A (no cases in women) |  | 0.66 (0.22 – 2.01) | 0.463 |
| Antihypertensive therapy (monotherapy vs. combination therapy) | 0.51 (0.22 – 2.00) | 0.123 | 0.67 (0.36 – 1.27) | 0.219 |
| Medication adherence | **0.49 (0.26 – 0.94)** | **0.031** | **0.48 (0.32 – 0.71)** | **<0.001** |

In the multivariable logistic regression models adjusted for medication adherence and duration of hypertension, the adjusted ORs of frailty on uncontrolled BP were 1.70 (1.00 – 2.90) in women, 0.84 (0.57 – 1.25) in men, women to men ratio of ORs 2.02 (1.04 – 3.92) (Table 3).

**Table 3.**
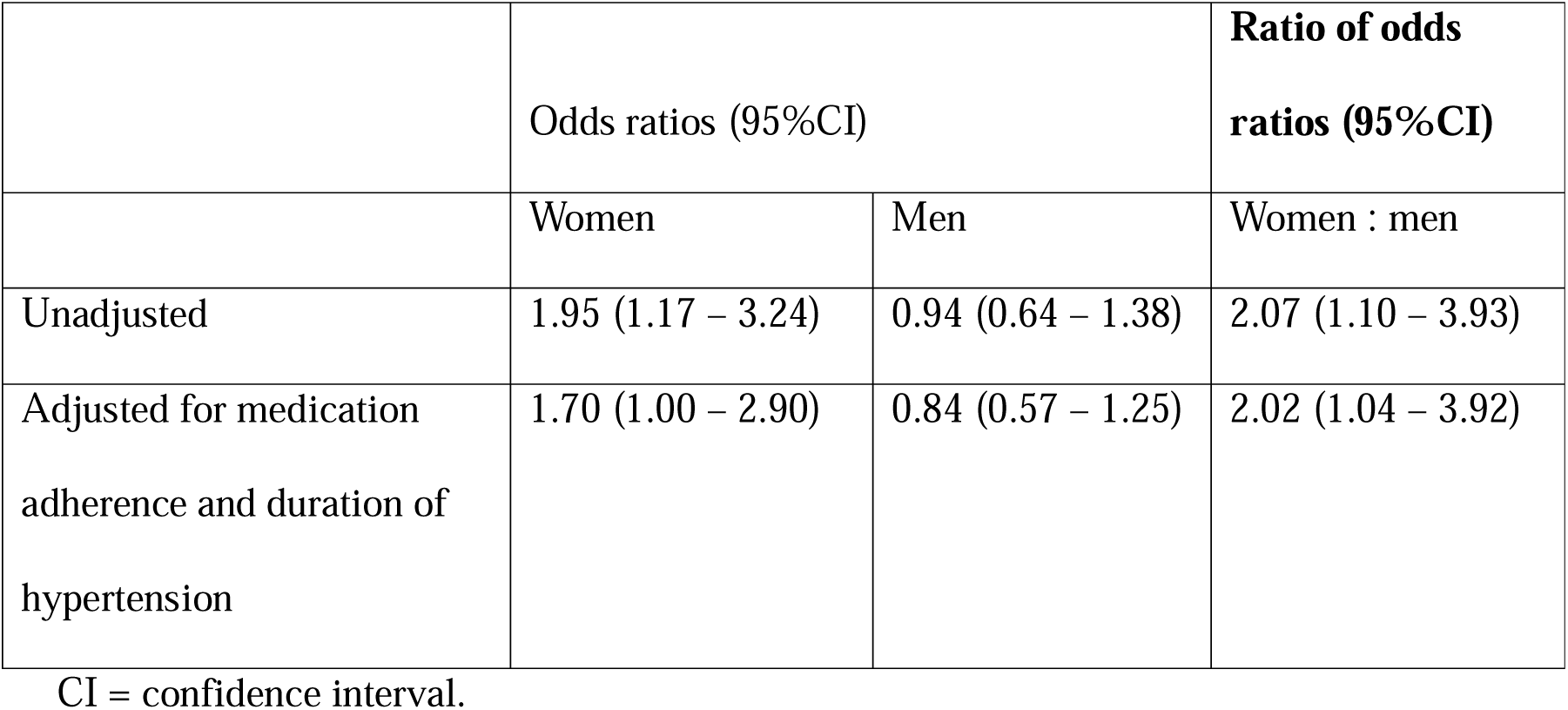
Associations between frailty and uncontrolled blood pressure.

## Discussion

In this study in participants with hypertension, we found that frailty was present in 28.6% of the participants and women had a significantly higher prevalence of frailty than men. Frailty was associated with an increased risk of having uncontrolled BP in women, with the effect being twice as strong in women compared to men.

Frailty is a common condition in older adults with cardiovascular disease and is associated with various adverse health outcomes.^33,34^ For older adults with hypertension, there is a greater risk of developing frailty which can impact overall care.^34^ The prevalence of frailty in our study population is similar to other studies in Vietnam in older adults with type 2 diabetes and coronary heart disease.^9,14^ However, there is limited evidence on sex differences in frailty in Vietnam. In a study of 638 older patients with diabetes (median age 71 years) attending outpatient clinics in Vietnam during 2019-2020, the prevalence of frailty defined by Fried’s frailty phenotype was 28.2% and there was no significant difference in frailty prevalence among women and men (29.9% vs. 26.8%, respectively, p=0.388). Differences in frailty definitions may explain this discrepancy.

There have been many studies on the prevalence of frailty in older people with hypertension, but limited studies have evaluated blood pressure control in older adults with frailty.^34,35^ Our findings are in line with a study on the association between frailty and hypertension treatment in Korea.^35^ In a study of 4352 older Koreans (mean age 72.6 years, SD 5.4), the proportion of participants with uncontrolled BP was higher in the frail (20%) compared to those who were prefrail (18%) and non-frail (15%), p=0.005.^35^

In our study, older women with frailty had the highest rate of uncontrolled hypertension. Frailty may have a greater impact on poor control of hypertension in women than in men due to various sex differences, including socioeconomic factors, lifestyles such as nutrition and physical exercises, different comorbidity patterns, and biological differences. These sex differences compound the effects of frailty on BP control. When comparing male and female participants in our study, we found that women were less likely to have higher education (31.0% of women vs 48.9% of men). The influence of educational background warrants further examination, particularly regarding its impact on health literacy, awareness of hypertension and medication adherence. The female participants in our study were also less likely to take part in regular exercise (27.9% vs 58.6% in men). While frailty can contribute to more sedentary lifestyles, the significant differences between women and men indicate a potential area for further investigation. In terms of comorbidities, there has been evidence that women are at a higher risk for dementia and cognitive decline compared to men,^36^ which can exacerbate the impact of frailty on BP control by reducing the ability to perform daily activities independently, including taking medications. Other comorbidities such as osteoporosis and falls can also impact the treatment of hypertension. In our study, the prevalence of falls and osteoporosis was higher in female participants compared to men. Since low blood pressure can sometimes increase the risk of falls and fracture, it is possible that physicians preferred higher BP targets for women to help manage this risk. In addition, sex differences in pharmacokinetics and pharmacodynamics can affect the efficacy of antihypertensive medications.^37,38^ In the ACCOMPLISH (Avoiding Cardiovascular Events through Combination Therapy in Patients Living with Systolic Hypertension) trial, which was designed to test the hypothesis that treatment with an angiotensin-converting enzyme (ACE) inhibitor combined with amlodipine would result in better cardiovascular outcomes than treatment with the same ACE inhibitor combined with a thiazide diuretic, the investigators found that benazepril – amlodipine combination was superior to the benazepril - hydrochlorothiazide combination in reducing cardiovascular events in patients with hypertension.^39^ However, subgroup analysis by sex revealed that the study results did not reach significance for women (hazard ratios for the primary outcome of cardiovascular events and cardiovascular death were 0.83, 95%CI 0.68 – 1.01, p = 0.06 in women vs. 0.80, 95%CI 0.69-0.91 in men, p=0.001).^39^

### Implications

Given the significance of sex differences on frailty and BP control, personalized treatment strategies should be explored when designing future strategies for improving BP management in the older population. A one-size-fits-all approach to hypertension management may not be effective because of the heterogeneity of this population. Special attention should be given to how frailty impacts older women and men differently. Understanding these factors can help in developing targeted interventions aimed at reducing frailty in women, such as strength training, improving access to healthcare, and enhancing nutrition programs. With the prefrail category (CFS 3) being the most common frailty category in both men and women in our study, it suggests that older adults with hypertension should be routinely screened for frailty to enable early detection of frailty. Timely intervention can help prevent the progression of frailty and its adverse effects on BP control, especially in older women.

### Strengths and limitations

To the best of our knowledge, this is the first study examining sex differences in how frailty affects BP control in older adults with hypertension. Our study provides important insights into the relationship between sex differences, frailty and BP control in older adults with hypertension. Despite the strength, there are potential limitations to consider. We did not collect data on socioeconomic status or nutrition intake, factors that may serve as potential confounders for the relationship between frailty and BP control. Future studies would benefit from including these parameters to provide a more comprehensive analysis and to enhance the robustness of the findings. Previous studies in adults with cardiovascular diseases in Vietnam reported that women had lower income compared to men.^40,41^ This disparity may lead to reduced access to healthcare and antihypertensive medications for women. Women may have caregiving roles, which might have limited their ability to engage in activities that promote physical activities and self-care. Furthermore, sex differences related to dietary intakes were not assessed in our study. In a study in 2203 community-dwelling adults in Vietnam, the authors found a stronger association between salt intake and hypertension in women compared to men.^42^ Finally, since the data collected in our study was tied to clinical care, future research involving laboratory data could deepen our understanding on sex differences in the impact of frailty on health outcomes in older adults. This knowledge could also aid in developing future care strategies that are specific to each gender.

## Conclusion

In this study in participants with hypertension, frailty was more common in women and was associated with an increased risk of having uncontrolled hypertension in women alone. This study contributes to the understanding of how frailty and sex influence blood pressure control in older adults with hypertension, and informs the development of targeted interventions to improve health outcomes for this population. These findings highlight the need for sex-specific approaches in managing blood pressure in older populations. Future studies in Vietnam should also examine sex differences in the impact of uncontrolled BP on cardiovascular outcomes in older adults.

## Data Availability

All data produced in the present study are available upon reasonable request to the authors.

